# Epidemic Persistence: Equilibria and Stability Analysis of Spread Process Dynamics over Networks, with Asymptomatic Carriers and Heterogeneous Model Parameters

**DOI:** 10.1101/2023.01.20.23284824

**Authors:** Xiaoqi Bi, Carolyn L. Beck

## Abstract

We present an analysis of epidemiological compartment models that explicitly capture the dynamics of asymptomatic but infectious individuals. Our models can be viewed as an extension to classic SIR models, to which a distinct Asymptomatic compartment is added. We discuss both a group compartment model capturing a Susceptible-Asymptomatic-Infected-Recovered-Susceptible (SAIRS) epidemic process, and also introduce and evaluate SAIRS dynamics evolving over networks. We investigate equilibria and stability properties that include both disease-free and endemic equilibria states for these models, providing sufficient conditions for convergence to these equilibria. Model parameter estimation results based on local test-site and Peoria county clinic data are given, and a number of simulations illustrating the effects of asymptomatic-infected individuals and network structure on the spread and/or persistence of the disease are presented.

## 1 Introduction

Modeling, analysis and control of epidemic spread processes over networks have been of interest in multiple communities over the past two decades, owing not only to the COVID-19 pandemic, but also to outbreaks of the related SARS and MERS viruses, Zika, Ebola, and more generally, computer network viruses and propogating opinions over social media networks. Conducting experiments to analyze infectious disease spread processes and response policies are prohibitive for many reasons, including not only costs, but more importantly ethics. As a result, mathematical modeling and simulation approaches provide essential alternatives for estimating and predicting when and how an epidemic might spread over a contact network [1]. Further, simulations of strategic control policies for validated epidemic models can provide insights into approaches for mitigating virus spread over networks [2].

The mathematical models for most epidemiological studies today derive from the *compartment models* first proposed by Kermack and McKendrick [3], although mathematical models for epidemics, or spread processes more generally, have been analyzed and studied for over 200 years, with one of the earliest known studies in the literature being that by D. Bernoulli on the analysis of the small-pox virus [4]. The now widely used compartment models assume every subject lies in a specific segment or compartment of the population at any given time, with these compartments including *susceptible* (S), *infected* (I), *exposed* (E) and/or *recovered* (R) population groups, leading to the classical epidemiological models: SI (susceptible-infected), SIS (susceptible-infected-susceptible), SIR (susceptible-infected-recovered) and SEIR (susceptible-exposed-infected-recovered) models. As one example, the Kermack and McKendrick SIS model is given by

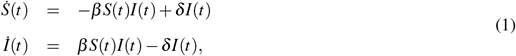

where *S*(*t*) is the susceptible (non-infected) segment of the population at time *t, I*(*t*) is the infected segment of the population at time *t, β* represents the rate of infection or contact amongst infected and susceptible subgroups, and *δ* represents the healing rate. This foundational model assumes: (1) a homogeneous population with no vital dynamics, that is birth and death processes are not included, meaning that infection and healing are assumed to occur at faster rates than vital dynamics and the population size is assumed to remain constant; and (2) the population mixes over a trivial network, or in other words, over a complete graph structure. These assumptions have led to errors in previous epidemic forecasts [5].

We note that similar models to that given in (1) have been derived for SI, SIR(S) and SEIR(S) processes; SI models simply have *δ* = 0; SIR(S) models include a recovered segment of the population and a recovery rate *γ*; and SEIR(S) models include an exposed segment of the population and a corresponding parameter *σ* capturing the rate at which an exposed individual transitions to the infected state; the exposed segment is typically assumed to be non-infectious with the accompanying rate parameter capturing the disease incubation period. There are numerous variants of these models, including recent models in which human awareness is taken into account [6–9], and in which multiple epidemic processes or epidemic processes with heterogeneous or non-static parameters may be propagating simultaneously [10–13].

Over the past two decades, both to address the discrepancies found in prior epidemic forecasts, and to better model spreading processes of computer viruses over communication networks, there has been an increased focus on the study of epidemic processes evolving over arbitrary network, or graph, structures; see for example [14–18], and from a controls perspective [19–21] (as the literature in this area is vast this list is not exhaustive). These networks represent the variation in interactions among members of a population, where the nodes in the network may represent either individuals or subgroups in the larger population, and the edges between nodes in the network represent the strength of the interaction between the nodes.

Over a network of *n* total nodes, epidemic or spread process dynamics can be described by Markov process models, for example, of dimension 2^*n*^ for SIS models and 3^*n*^ for SIR models. These models describe the probability of each node transitioning from susceptible to infected, and/or to recovered states, and back, where the probabilities are determined by the model rate parameters (infection, healing, etc.) and the network interconnection structure, and reflect the stochastic evolution of such epidemic processes. Clearly, as the number *n* of nodes in the network increase, analysis of these models becomes intractable. As an alternative, *mean-field approximation* (MFA) models have been derived and shown to be appropriate under certain assumptions; these models are derived by taking expectations over infection transition rates of the agents and rely on the fundamental work of Feller [22] and Kurtz [23].

When individuals or population subgroups are assumed to be interconnected via a graph with adjacency matrix *W* = [*W*_*i j*_],where element *W*_*i j*_ defines the strength of the connection from node *i* to node *j*, and further making assumptions of large and constant agent population size and probabilistic independence assumptions, the deterministic networked MFA dynamic models are now considered standard models; these models have been analyzed in detail and shown to provide upper bounds on the probability of infection of a given agent at any given time (see [24] and [25] for discussions and perspectives). Again considering an SIS process example, denoting the probability of node *i* being infected at time *t* by *p*_*i*_(*t*) ∈ [0, 1], the following differential equation provides a MFA model of the evolution of the probabilities of infection of the nodes:

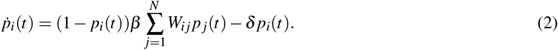

This model provides a lower complexity deterministic approximation to the full dimension Markov process model of a SIS spread process evolving over a static network. Further details can be found in [19, 26, 27]. Discrete time versions of these approximation models have also been proposed and studied, see for example [28, 29].

The main objectives in most analyses of epidemic process dynamics include computing the system equilibria, and determining the convergence behavior of these processes near the equilibria. In particular, conditions for the existence of and convergence to “disease-free” or “endemic” equilibria are sought. For (2), it is straightforward to see that the disease-free state, 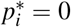 for all *i* ∈ {1,…, *N*}, is a trivial equilibrium of the dynamics. It has been shown that this equilibrium is globally asymptotically stable if and only if 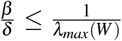, where *λ*_*max*_(*W*) represents the largest real-valued part of the eigenvalues of the matrix *W*. It has further been shown, however, that if 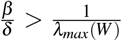, then there exists another equilibrium that is (almost) globally asymptotically stable, with 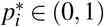 for all *i* ∈ {1, 2, …, *n*}, implying the system converges asymptotically to an *endemic* state [28, 30–32].

In this paper we consider a compartment model structure that specifically accounts for *infectious but asymptomatic* subgroups or individuals, namely a SAIRS model structure, incorporating Susceptible(S), Asymptomatic-infected(A), Infected-symptomatic(I), and Recovered(R) subsets of the population. We note that the asymptomatic subset we consider may include those individuals who do not experience symptoms through the course of their infection, as well as pre-symptomatic individuals. This structure may be used to directly capture the dynamics of COVID-19 and the role asymptomatic individuals play in the disease spread process; this model was first introduced in public online seminars and panel discussions in [33, 34], and in the literature in [20, 35]. Compartment models with different structures but including explicit asymptomatic population segments were previously proposed for dengue fever [36] and rumor spreading over online social networks [37]. Relevant work on alternative SAIRS model structures has been reported in [38–41]. In [38], the authors derive mean-field approximations of the exact state evolution for SAIRS models, and also present a game-theoretic model where nodes choose their activation probabilities in a strategic (e.g., selfish) manner using current state information as feedback. The author in [39] introduces a compartmental model including a group of individuals with pure asymptomatic infection (i.e., having no symptoms throughout the course of infection), with permanent immunity upon recovery, and provides estimations of the asymptomatic populations in California, Florida, New York, and Texas. The authors in [40] present a more complex data-informed model including pre-symptomatic, asymptomatic, and hospitalized subgroups of the population, and provide forecasts for the epidemic over homogeneous populations. In [41] the authors provide ℋ_∞_ based (i.e., worst-case) stability analyses for an SAIR model structure, and provide more focused simulations for SAIR spread processes over small-world networks; there is no explicit loss of immunity included in their model under which to study endemicity behavior. Herein we provide more thorough stability analyses and simulations of the endemic equilibria for SAIRS models (as well as for the disease free equilibria), than have been presented in prior work. Our analysis approach is based on classic Lyapunov methods for dynamical systems. We further take the contact network into consideration and discuss the impact of the network structure and potentially heterogeneous epidemic parameters on the spread process.

In the remainder of the paper, we first present the specific SAIRS group and networked models we will consider throughout, and discuss the equilibria and stability properties of these models in Section 2. In Section 3, we discuss a simple least squares estimation approach to compute the SAIRS model parameters from data, which relies on knowledge of the proportion, *q*, of the infections that are asymptomatic. We therefore also discuss methods for estimating this proportion, and use local COVID test-site data (Champaign County Public Health District) to evaluate the results. These initial estimation results are compared to data recorded at Peoria County clinics from April 2020 to July 2020, which explicitly includes symptoms of all sample individuals. We then discuss a series of simulation studies in Section 4, which illustrate our stability results as well as highlighting the role the asymptomatic subgroup and the contact network play in disease spread under various quarantine policies made with and without awareness of asymptomatic status. We further present a longer-term forecast for the epidemic process with both pharmaceutical and non-pharmaceutical mitigation approaches. To conclude, we discuss the challenges the currently available data present and our ongoing and future work in Section 5.

## 2 The SAIRS model

In order to investigate the effects of asymptomatic individuals on the spread of the epidemic, we consider the effects of a proportion of the infected subgroup being asymptomatic and potentially unaware of their carrier status. We evaluate both single group models as well as networked models, providing equilibria and stability analyses.

### 2.1 Single-Group and Networked Models

Let *S*(*t*), *A*(*t*), *I*(*t*), *R*(*t*), respectively, represent the proportion of susceptible, asymptomatic-infected, symptomatic-infected, and recovered individuals at time *t*. The Group SAIR(S) model we consider is characterized

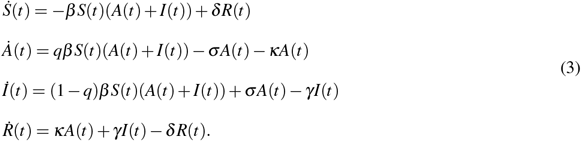

Here again *β* is the transmission rate amongst susceptible and infected groups, the latter of which includes both asymptomatic and symptomatic; *κ* and *γ*, respectively, are the recovery rates for asymptomatic-infected and symptomatic-infected groups. The proportion of infections that are asymptomatic (and/or pre-symptomatic) is denoted by *q*, after which the newly infected individuals show no symptom but are still infectious; correspondingly, (1 − *q*) represents the proportion of symptomatic infections. Further, *σ* is the progression rate from asymptomatic (and/or pre-symptomatic) to symptomatic, and *δ* represents the rate at which immunity recedes. When *δ* = 0, individuals gain permanent immunity to the infection upon recovery. We assume these relations hold for all *t* ≥ 0.

We also study the SAIRS model dynamics of *n* agents (individuals or subpopulations) interconnected over an arbitrary network structure, with adjacency matrix denoted by *W*. Defining *s*_*i*_, *a*_*i*_, *p*_*i*_, *r*_*i*_, respectively, as the proportion of the subpopulation *i* that is susceptible (or healthy), asymptomatic-infected, symptomatic-infected, or recovered, the Networked SAIRS (N-SAIRS) dynamics over an arbitrary interconnection network is given by

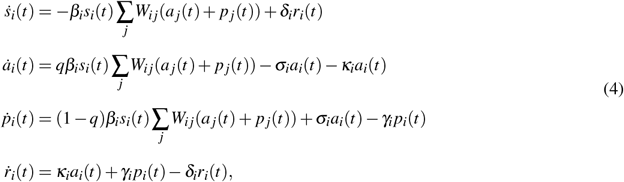

where, similar to the Group Model (3), for a subpopulation *i, β*_*i*_ is the agent-to-agent transmission rate; *κ*_*i*_ and *γ*_*i*_, respectively, are the recovery rates for asymptomatic-infected and symptomatic-infected subsets; again, *σ*_*i*_ represents the transition rate from asymptomatic to symptomatic infected; and *δ*_*i*_ represents the rate at which individuals may be susceptible to reinfection again after recovery. Since all individuals in a subgroup *i* will reside in one of these subsets, we have *s*_*i*_(*t*) + *a*_*i*_(*t*) + *p*_*i*_(*t*) + *r*_*i*_(*t*) = 1, over all *i* ∈ [*n*]. This proportion is relative to the subpopulation size, *N*_*i*_ of group *i*; recall the total population *N* = ∑_*i*_ *N*_*i*_.

#### Remark

In the case where we have homogeneous spread parameters and the underlying network topology is complete with evenly distributed interconnection weights, that is, when *W*_*i j*_ = 1*/n* for all *i, j* ∈ [*n*], and (*β*_*i*_, *κ*_*i*_, *γ*_*i*_, *σ*_*i*_, *δ*_*i*_) = (*β, κ, γ, σ, δ*) for all *i* ∈ [*n*], the Group Model (3) and the Networked Model (4) are equivalent.

Prior to discussing the analysis of equilibria and stability for these models, we note the following result which establishes that the N-SAIRS model is well-defined. This result was first presented in [20] for the discrete-time case using an induction argument; it is straightforward to adapt this result to the continuous-time model given in (4). We first state our assumption on the model parameters.

#### Assumption 1.

*For all i, j* ∈ [*n*], *we have β*_*i*_, *γ*_*i*_, *δ*_*i*_, *σ*_*i*_, *δ*_*i*_, *W*_*i j*_ *≥* 0, 0 ≤ *q ≤* 1.

#### Lemma 1.

*Consider the model in (4) under Assumption 1. Suppose s*_*i*_(0), *a*_*i*_(0), *p*_*i*_(0), *r*_*i*_(0) ∈ [0, 1], *s*_*i*_(0) + *a*_*i*_(0) + *p*_*i*_(0) + *r*_*i*_(0) = 1, ∀*i* ∈ [*n*]. *Then, for all t* ≥ 0 *and i* ∈ [*n*], *we have s*_*i*_(*t*), *a*_*i*_(*t*), *p*_*i*_(*t*), *r*_*i*_(*t*) ∈ [0, 1] *and s*_*i*_(*t*) + *a*_*i*_(*t*) + *p*_*i*_(*t*) + *r*_*i*_(*t*) = 1.

*Proof:* We show that for all *i* ∈ [*n*] and *t* ≥ 0, when *s*_*i*_(*t*) = 0, *a*_*i*_(*t*) = 0, *p*_*i*_(*t*) = 0, *r*_*i*_(*t*) = 0, respectively, we have 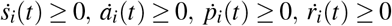; and when *s*_*i*_(*t*) = 1, *a*_*i*_(*t*) = 1, *p*_*i*_(*t*) = 1, *r*_*i*_(*t*) = 1, respectively, we have 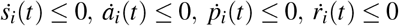.

Firstly, from *s*_*i*_(0) + *a*_*i*_(0) + *p*_*i*_(0) + *r*_*i*_(0) = 1, and 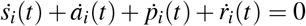, we have *s*_*i*_(*t*) + *a*_*i*_(*t*) + *p*_*i*_(*t*) + *r*_*i*_(*t*) = 1, ∀*i* ∈ [*n*], ∀*t* ≥ 0.

By Assumption 1 and (4), for all *i* ∈ [*n*], if *s*_*i*_(0) = 0, we have 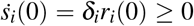. By the continuity of *s*_*i*_(*t*), there exists 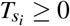, such that, 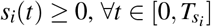. Similarly, if *a*_*i*_(0) = 0, we have 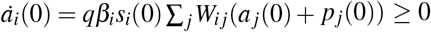; if *p*_*i*_(0) = 0, we have 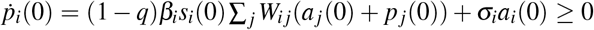; and if *r*_*i*_(0) = 0, we have 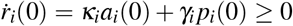. Thus, there also exist 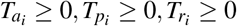, respectively, such that 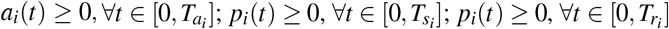.

Define 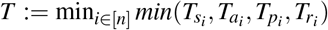 for *i* ∈ [*n*]. Then by definition, *s*_*i*_(*T*) *≥* 0, *a*_*i*_(*T*) *≥* 0, *p*_*i*_(*T*) *≥* 0, *r*_*i*_(*T*) *≥* 0, ∀*i* ∈ [*n*]. Similarly, we have 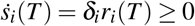 if *s*_*i*_(*T*) =0; 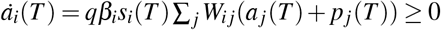 if *a*_*i*_(*T*)= 0; 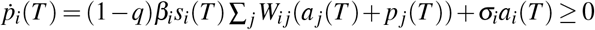 if *p*_*i*_(*T*)=0; 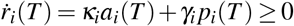 if *r*_*i*_(*T*)=0. Thus, for all *t* ≥ 0 such that *s*_*i*_(*t*) = 0, *a*_*i*_(*t*) = 0, *p*_*i*_(*t*) = 0 or *r*_*i*_(*t*) = 0, respectively, we have 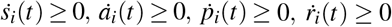. This further suggests that, *s*_*i*_(*t*) *≥* 0, *a*_*i*_(*t*) *≥* 0, *p*_*i*_(*t*) *≥* 0, *r*_*i*_(*t*) *≥* 0, ∀*i* ∈ [*n*], ∀*t* ≥ 0.

Next, we prove that if *s*_*i*_(*t*) = 1, *a*_*i*_(*t*) = 1, *p*_*i*_(*t*) = 1, or *r*_*i*_(*t*) = 1, respectively, we have 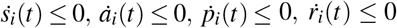. Given in Lemma 1, *s*_*i*_(*t*) + *a*_*i*_(*t*) + *p*_*i*_(*t*) + *r*_*i*_(*t*) = 1, and *s*_*i*_(*t*), *a*_*i*_(*t*), *p*_*i*_(*t*), *r*_*i*_(*t*) *≥* 0, ∀*i* ∈ [*n*]. Hence, if *s*_*i*_(*t*) = 1, we have *a*_*i*_(*t*) = 0, *p*_*i*_(*t*) = 0, *r*_*i*_(*t*) = 0, which leads to 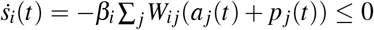. Similarly, if *a*_*i*_(*t*) = 1, we have 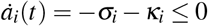; if *p*_*i*_(*t*) = 1, 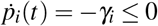; and if 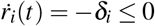. Similar to the preceding argument, we have *s*_*i*_(*t*) ≤ 1, *a*_*i*_(*t*) ≤ 1, *p*_*i*_(*t*) ≤ 1, *r*_*i*_(*t*) ≤ 1, ∀*i* ∈ [*n*], ∀*t* ≥ 0.

### 2.2 Equilibria and stability

To quantitatively and qualitatively evaluate the propagation of the virus, a critical threshold quantity, denoted by *R*_0_ and referred to as the basic reproduction number, is used extensively in epidemiological studies. This number indicates how rapidly infected individuals transmit the virus to healthy individuals. In this section, we evaluate the SAIRS model equilibria and conduct stability analyses around the equilibria, leading to conditions on *R*_0_ which provide quantitative criteria for convergence to the disease-free state, or to an endemic state. We first consider the group model.

#### 2.2.1 Group Model SAIRS

Noting that *S*(*t*) = 1 −*A*(*t*) −*I*(*t*) −*R*(*t*), the nonlinear system dynamics (3) can be written as

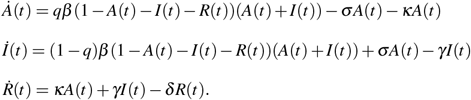

By setting 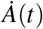, *İ*(*t*), 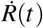 to 0, we can see immediately that an equilibrium state of system (5) is given by (*A*^*e*^, *I*^*e*^, *R*^*e*^) = (0, 0, 0) with *S*^*e*^ = 1. This is the disease-free equlibrium (DFE) in the case of non-permanent immunity. Linearizing system (5) around (*A*^*e*^, *I*^*e*^, *R*^*e*^), we obtain the system Jacobian matrix,

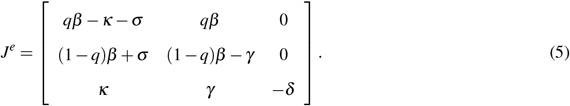

The system described by (5) is globally asymptotically stable around the DFE if all eigenvalues of *J*^*e*^ have negative real parts; see Theorem 4.7 from [42]. Computing the characteristic polynomial for *J*^*e*^, we have after some straightforward manipulations,

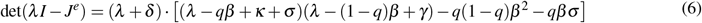

Applying the Routh-Hurwitz criterion to (6) gives us the following.

##### Proposition 1.

*Given the system dynamics defined by (5), the DFE* (*S*^*e*^, *A*^*e*^, *I*^*e*^, *R*^*e*^) = (1, 0, 0, 0) *is globally asymptotically stable (GAS) when δ* > 0 *and*

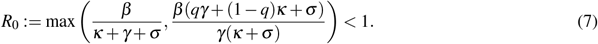

In the case where *δ* = 0, that is when immunity following recovery from infection is permanent, the disease-free equilibria will be the subspace of points (*S*^*e*^, *A*^*e*^, *I*^*e*^, *R*^*e*^) = (*c*_*S*_, 0, 0, *c*_*R*_), where constants *c*_*R*_, *c*_*S*_ satisfy *c*_*S*_ + *c*_*R*_ = 1. Analyzing the Jacobian for (5) in this case gives us that the equilibria (*S*^*e*^, *A*^*e*^, *I*^*e*^, *R*^*e*^) = (*c*_*S*_, 0, 0, *c*_*R*_) are also globally asymptotically stable (GAS) when (7) is satisfied. That is, this basic reproduction number expression provides an appropriate threshold for determining when the spread process for the SAIRS model will or will not spread exponentially in either of the scenarios of permanent or non-permanent immunity.

We may also consider the case where the asymptomatic-infected and symptomatic-infected individuals have different infection transmission rates. In the case of COVID-19, this difference could be partly due to the inability to conduct frequent large-scale population testing, for example allowing efficient identification and isolation of Asymptomatic individuals. Thus, we would have different quarantine control effectiveness over these two subpopulations. In this case, we denote the infection transmission rates for agent-to-agent contact between the susceptible subgroup and the two infectious groups, respectively, as *β*_*A*_, *β*_*I*_. As in the preceding analysis, we compute the Jacobian around the disease-free equilibrium (*S*^*e*^, *A*^*e*^, *I*^*e*^, *R*^*e*^) = (1, 0, 0, 0), as

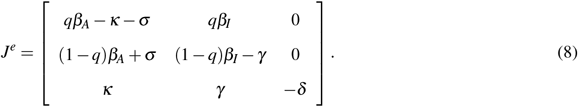

Following a similar approach as before yields

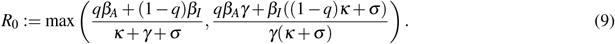

For GAS, using a similar argument, we can again show it is required that *R*_0_ *<* 1.

Of perhaps greater interest is the endemic equilibria for (5). If we again assume non-permanent immunity, that is, *δ* > 0, setting 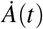, *İ*(*t*), 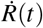 to 0, we can compute the unique endemic equilibrium for (5):

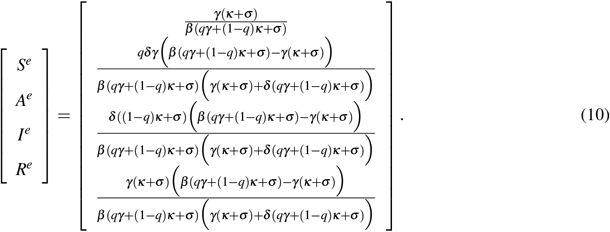

Denoting Ψ = *γ*(*κ* + *σ*) and Φ = *qγ* + (1 − *q*)*κ* + *σ*, and noting that both Ψ > 0 and Φ > 0, we further define

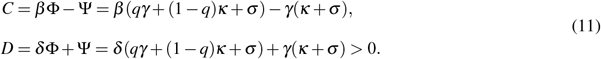

The endemic equilibrium now can be written more compactly as

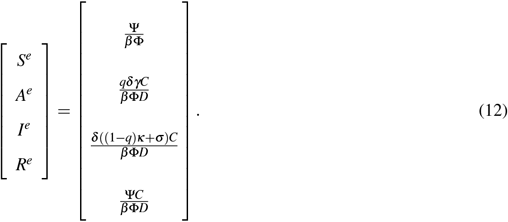

Using the relationship *S*(*t*) = 1 − *A*(*t*) − *I*(*t*) − *R*(*t*) and the expression for the endemic equilibrium point in (12), we compute the Jacobian around this equilibrium point as

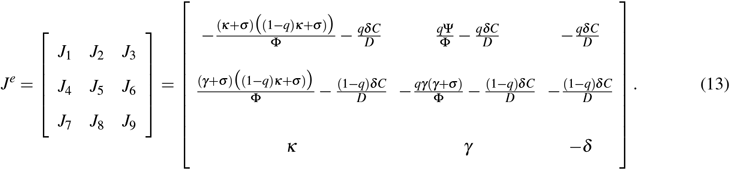

We can then show the following stability result using the Routh-Hurwitz criterion and algebraic manipulations.

##### Proposition 2.

*Given the system with dynamics defined by (5), with endemic equilibrium (12), and C and D as defined in (11) and (13) respectively, if both C* > 0 *and*

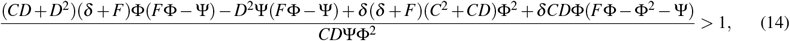

*where F* = *γ* + *κ* + *σ, then the system asymptotically converges to the endemic equilibrium*.

*Proof:* Note that the characteristic polynomial for *J*^*e*^ is given by

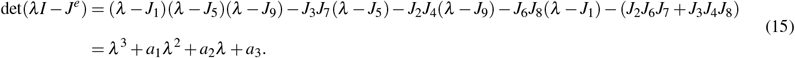

Applying the Routh-Hurwitz criteria to (15), for all roots of the polynomial to have negative real parts, we require *a*_1_, *a*_3_ > 0, and *a*_1_*a*_2_ > *a*_3_. Assuming

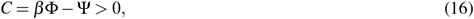

we have, *J*_1_, *J*_5_ *<* 0, *J*_3_, *J*_6_, *J*_9_ ≤ 0, and *J*_7_, *J*_8_ *≥* 0. Consequently,

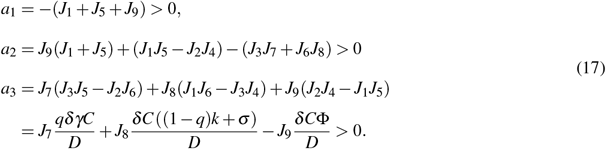

To satisfy the condition *a*_1_*a*_2_ > *a*_3_, we equivalently require

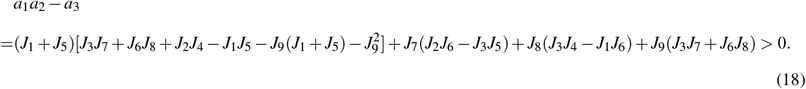

After many tedious algebraic manipulations we can show that satisfying (18) is equivalent to satisfying the condition

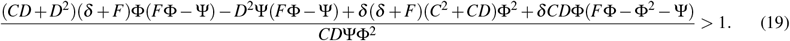

Inequalities (16) and (19) provide a sufficient condition for asymptotic stability of the endemic equilibrium. Simulations illustrating the behavior of the SAIRS model dynamics when conditions for asymptotic stability to the endemic equilibrium are met are given in Section 4.

#### 2.2.2 Networked Model N-SAIRS

We now evaluate equilibria and their stability properties for the networked SAIRS models. Given *s*_*i*_(*t*) = 1 − *a*_*i*_(*t*) − *p*_*i*_(*t*) −*r*_*i*_(*t*) for all *t* ≥ 0, *i* ∈ [*n*], system (4) can be represented in matrix form as

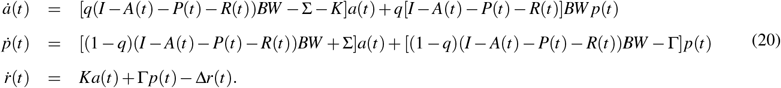

Here,

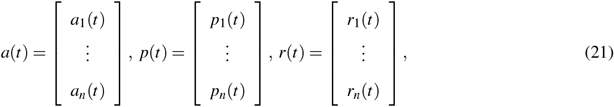

with *n × n* matrices *A*(*t*) = *diag*(*a*_*i*_(*t*)), *P*(*t*) = *diag*(*p*_*i*_(*t*)), *R*(*t*) = *diag*(*r*_*i*_(*t*)), *B* = *diag*(*β*_*i*_), *K* = *diag*(*κ*_*i*_), Γ = *diag*(*γ*_*i*_), ∑ = *diag*(*σ*_*i*_), Δ = *diag*(*δ*_*i*_), and adjacency matrix *W*.

We first consider the case with permanent immunity, i.e., *δ*_*i*_ = 0. Setting 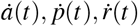 to 0, we can compute the equilibrium state where 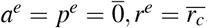, where 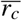 is a non-negative constant vector with elements 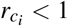. Linearizing the system (20) at the equilibrium (*a*^*e*^, *p*^*e*^, *r*^*e*^), we obtain the 3*n* × 3*n* system Jacobian Matrix given by

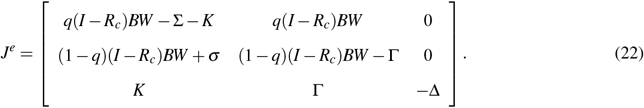

Analysis of this Jacobian matrix will lead to a constraint on the spectrum of the weighting matrix *W*, which if met guarantees the system is at least locally asymptotically stable at the DFE. An alternative is to consider a Lyapunov stability analysis approach, which may provide global results. Specifically, if we consider a quadratic Lyapunov function, we can show the following.

##### Theorem 1.

*For the system given by (20), under Assumption 1, the DFE* 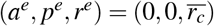 *is globally asymptotically stable (GAS) if*

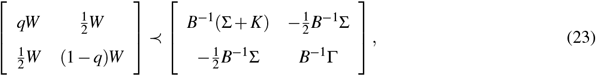

*where ≺ denotes relative definiteness of the matrices*.

*Proof:* We now consider non-permanent immunity (Δ > 0), thus *r*_*c*_ = 0. Consider the Lyapunov function

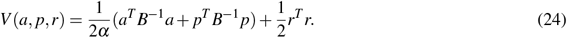

Clearly, *V* > 0 for all (*a, p, r*) ≠ 0 and

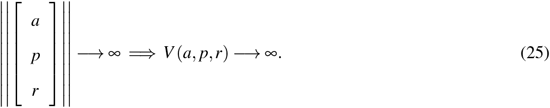

Computing the derivative, we have

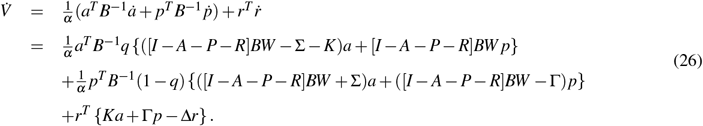

Since *A, P* and *R* are diagonal matrices with 0 ≤ *a*_*i*_ ≤ 1, 0 ≤ *p* _*j*_ ≤ 1, and 0 ≤ *r*_*k*_ ≤ 1 for all *i, j, k* = 1, …, *n*, and we know all elements of ∑, *K*, Γ and Δ are non-negative, then it is straightforward to see

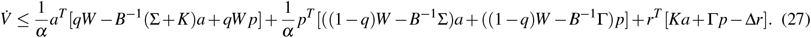

For GAS, we require 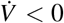 for all *t* ≥ 0. We note that the first two terms on the right hand side of (27) have no dependence on *r*; thus if these two terms together are negative, then by an appropriate selection of *α* we can always make 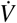 negative. We therefore can simplify the analysis by considering

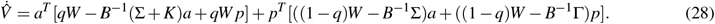

Applying a completion of squares and algebraic simplifications we can show the condition 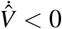 to be equivalent to the inequality

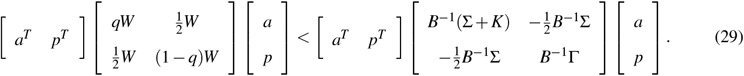

In the case where 1 ≥ *r*_*c*_ > 0, we can apply a basic translation. The result then follows from Theorem 4.2 in [42]. Note that directly from (27), the condition in (20) is also sufficient for GAS in cases with permanent immunity (Δ = 0). ◼

Summarizing, (23) provides a test that bounds the maximum eigenvalue of the *q*-scaled adjacency matrix *W* in terms of the minimum eigenvalue of a matrix consisting of diagonal block entries of ratios of healing and transition rates (*κ*_*i*_, *γ*_*i*_ and *σ*_*i*_) to infection rates (*β*_*i*_). This condition generalizes the usual *R*_0_ threshold to allow for heterogeneous infection parameters over multiple infection compartments in the N-SAIRS model form.

##### Remark 1.

*Note that in the case of a slightly simpler spread process model, for example for a networked SIRS model, a sufficient condition for convergence to the DFE would be λ*_max_(*W*) *< λ*_min_(*B*^−1^Γ) = min_*i*_(*γ*_*i*_*/β*_*i*_) = min_*i*_(1*/R*_*Oi*_).

## 3 Parameter estimation

In this section we discuss a simple least-squares approach for model parameter estimation for a discrete-time N-SAIRS model, given below in (30), and present some of our initial estimation results from local data for COVID-19. We also provide an overview of the approach we have used to estimate asymptomatic proportions of the subpopulations of interest.

The data sets we consider result from sampling on a daily basis, thus a discrete-time model is better suited for estimating and evaluating model parameters. We first apply a forward Euler’s method to the continuous-time networked model in (20), and appropriately scale the model by *N*_*i*_ for each subpopulation, giving us the discrete-time N-SAIRS model,

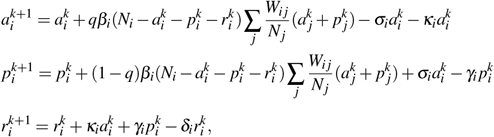

where {*a*_*i*_, *p*_*i*_, *r*_*i*_} represent the population of asymptomatic, symptomatic-infected and recovered individuals in region *i* respectively, ∀*i* ∈ [*n*]. Since our simulation update also will be daily and the sampling rate is once-per-day, the sampling parameter typically made explicit in such sampled-data models will be 1 and thus is not explicitly noted above.

### 3.1 Least Squares Estimation

When the proportion *q* of the asymptomatic infections is known or estimated, we can apply a simple least squares (LS) approach, for example as outlined in [43] and further described for SAIRS models in [20], to estimate the model parameters *β*_*i*_, *σ*_*i*_, *κ*_*i*_, *γ*_*i*_, and *δ*_*i*_. Our initial estimation step is therefore to estimate *q*.

#### 3.1.1 Estimating the Asymptomatic Population Proportion

Due to the difficulties in identifying and monitoring infected individuals without symptoms, explicit and unbiased information for asymptomatic-infected estimations is not always available. We have applied Nesterov’s Next-Day Law to estimate the daily number of asymptomatic individuals, based on a constant latent infectious period assumption, and further to estimate the proportion *q* of the asymptomatic infections as a fraction of the total population. We note that, more precisely stated, this approach gives us a pre-symptomatic subpopulation proportion. We state the Next-Day Law here for completeness.

##### Proposition 3.

*[44] Let T*(*d*) *represent the total number of confirmed cases by day d, and A*(*d*) *represent the number of asymptomatic infected individuals at the beginning of day d. Assume the latent period (the time between exposure and onset of symptoms) is a constant time of* Δ *days. Then, A*(*d* + 1) = *T* (*d* + Δ) −*T* (*d*), ∀*d* ∈ *ℤ*

From estimated daily asymptomatic numbers, the proportions *q* and 1 − *q* corresponding to the asymptomatic and symptomatic infections, can be approximated.

As observed in many data sets for COVID-19 (for example, in the testing data posted by the Champaign-Urbana Public Health Department, Illinois), the infected (both asymptomatic and symptomatic) and recovered populations are relatively much smaller than the susceptible population. Therefore, the third and fourth terms in the first two equations in (30) are assumed to be negligible compared to the second term. Omitting these terms gives us the approximate relationship

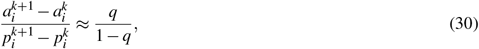

from which we can approximate

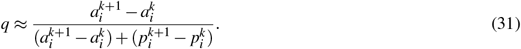

#### 3.1.2 Estimation of Model Parameters

Given an estimated, or a known value, for *q*, we can now rewrite the networked system (30) as a system of linear equations. Let

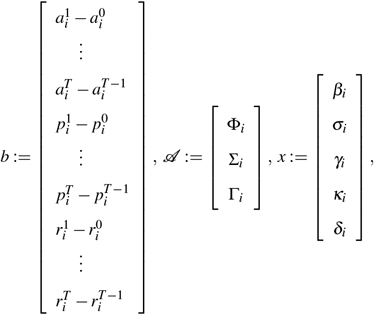

with

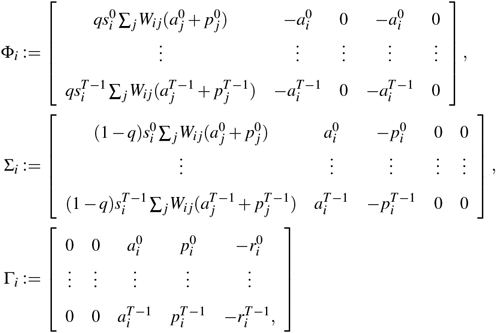

where 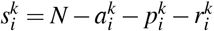, *k* ∈ *ℤ*.

Then the discrete-time N-SAIRS model can be written as

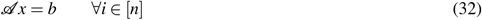

Since *q* is assumed known, (32) is linear with respect to the remaining model parameters. When *A* is full rank, we can thus recover the parameters 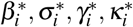, and 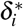 using a standard least-squares solution to (32).

### 3.2 Preliminary estimation results

For our preliminary parameter estimation, local COVID-19 testing-site data from Champaign County, Illinois, dating from April to September, 2020, is used. We obtained data from the Champaign-Urbana Public Health District website (publicly available at https://www.c-uphd.org/champaign-urbana-illinois-coronavirus-information.html), which is updated daily and includes the total accumulative number of infected (lab-confirmed), recovered, hospitalized, and deceased individuals for Champaign county, as well as current number of actively infected (lab-confirmed) individuals over 34 zip code areas within the county. We scraped the website data on a daily basis manually from April 2020 to March 2022, storing the daily accumulative infected, actively-infected, and recovered populations for each zip code in a Google Sheet [45]. Preliminary estimates are presented from the initial wave of COVID-19 in Champaign County, where we consider different phases of the epidemic according to the Illinois State Restore plan, specifically:

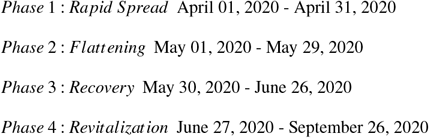

We assume a latent period of Δ = 6 days, giving estimated parameter values:

| Phases | $q$ | $\beta$ | $\sigma$ | $\gamma$ | $\kappa$ | $R_0$ |
| --- | --- | --- | --- | --- | --- | --- |
| Phase 2 | 0.7 | 0.06 | 0.22 | 0.15 | -0.10 | 1.004 |
| Phase 3 | 0.6 | 0.07 | 0.15 | 0.15 | -0.05 | 1.156 |
| Phase 4 | 0.6 | 0.07 | 0.08 | 0.11 | 0.02 | 1.104 |

Estimated local Sars-CoV-2 paramters

For comparison, we computed an asymptomatic infection proportion *q* using Peoria County medical clinics data [46], containing COVID-19-related records of all individuals who visited one of eight medical clinics in Peoria and Pekin from April to June 2020. The Peoria clinic data explicitly includes records of the COVID-19 test results, as well as all symptoms described by the patient to the provider, for each visiting individual. The computed values for proportion *q* for Phase 2 and Phase 3 in the Peoria data are approximately 0.51 and 0.42, respectively, both of which are lower than the values estimated from the Champaign County data. This difference is not unexpected, given that patients visited the Peoria clinics because they were experiencing symptoms of illness, although these may have been illnesses other than COVID-19.

We note that, as the epidemic progresses, the basic reproduction number *R*_0_ first increases, and then decreases due to the implementation of quarantine and other social distancing measures. The prelimimary results also expose issues with data-based estimation and analysis early in an epidemic. For example, due to the limited available testing in the early stage of the epidemic we have a non-random population sample, thus the testing population presented in the data is skewed toward Symptomatic-Infected individuals. This hinders us from accurately capturing the true proportion of the Asymptomatic-Infected subgroup, as well as an accurate prevalence of the epidemic over the total population [47]. In addition, our assumption of a constant latent period is not consistent with viral infections, including COVID-19; the latent period value we have chosen is thus an average value taken from [48, 49]. These issues lead to estimation errors, in particular note the negative values for recovery rate *κ* in Phase 1 and Phase 2. For this reason, estimated values from the larger virology and epidemiology literature are evaluated and used in our simulation studies.

## 4 Simulations

In this section, we illustrate the dynamics associated with endemicity, and the roles of the asymptomatic subgroup and contact network in the progression of the epidemic. We follow this with a multi-stage forecast of an epidemic spread process with both pharmaceutical and non-pharmaceutical mitigation approaches, as well as a simulation of an endemic COVID-19 process under annual vaccinations.

First, we simulate a baseline N-SAIRS model based on (4), for which we assume homogeneous spread parameters and a five-subpopulation network structure. We assume the total population size is 10, 000 and the respective subpopulations denoted A, B, C, D, and E have populations of 2000, 2500, 1500, 3500, and 500 people, respectively. In this baseline simulation we assume the subpopulations are fully connected with evenly distributed edge weights, thus this model is equivalent to the single group model represented in (3). We use the estimation results from early local data (discussed in Section 3) in addition to drawing upon the literature on COVID-19 (e.g., [48, 50–52]) to inform our baseline model parameter value selection, specifically setting (*q, β, σ, γ, κ, δ*) = (0.7, 0.25, 0.15, 0.11, 0.08, 0.0001).

These values represent the original strain of SARS-CoV-2. Note these parameters roughly correspond to an infectious disease with a duration of symptomatic infection of 9 days, duration of asymptomatic infection of 12 days, duration of pre-symptomatic infection of 6 days, and duration of immunity following recovery from infection of 30 years, or essentially permanent immunity. We further set the initial proportions of the *A, I, R* compartments as

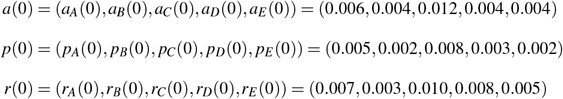

Simulating the SAIRS model over 60 days results in the epidemic progression shown in Fig 1. Note that peak active infection occurs on day 33, that is *p*(*t*) + *a*(*t*) attains a maximum of approximately 28% on day *t* = 33. By day 60, approximately 87% of the entire population has been or is infected; assuming a mortality rate of 2% would correspond to 174 deaths in the two month time span. Again we note this model assumes homogeneous mixing within the entire population. We will use this baseline model to compare to situations where immunity following recovery is not permanent, leading to endemicity and to potential virus mutations yielding multi-strain/multi-stage viral processes [53].

**Figure 1:**
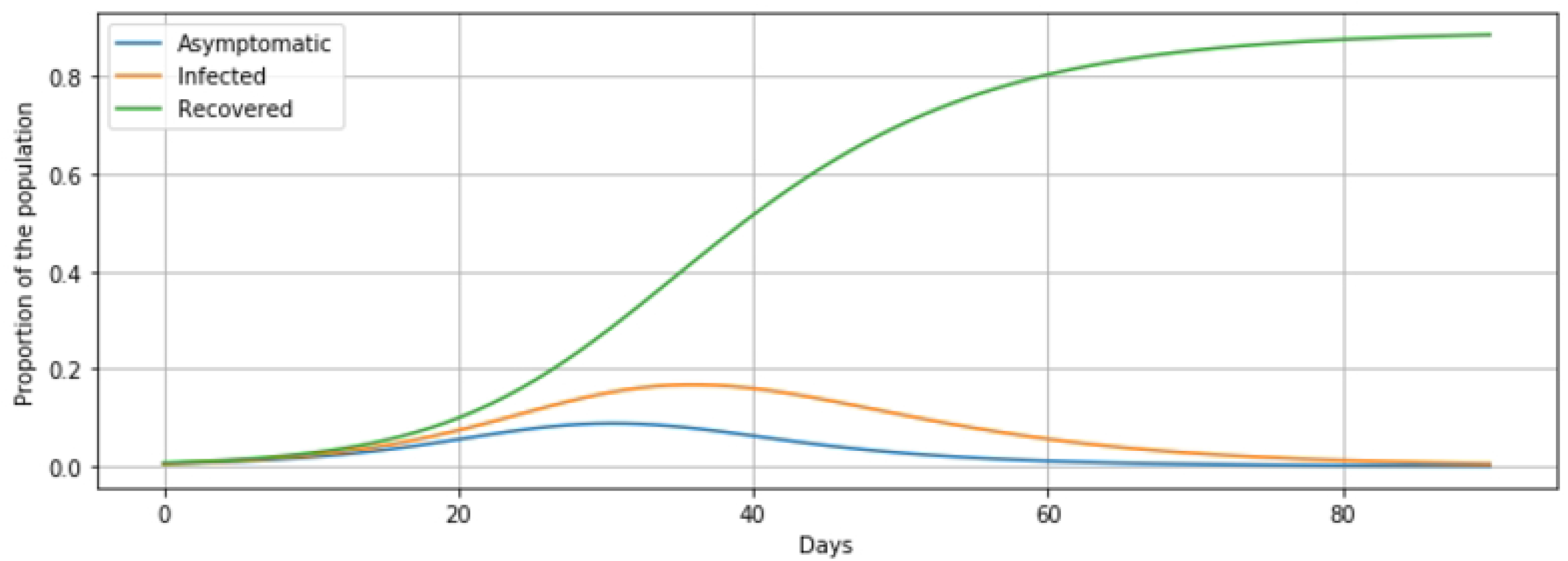
Group/Network SAIRS Simulation: Baseline Model

### 4.1 Endemicity

As presented in Proposition 2, the condition for endemicity (that is, GAS of the endemic equilibrium) is given by

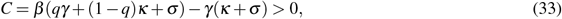

and

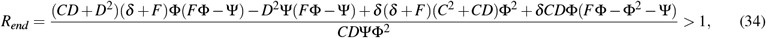

where *D, F*, Φ and Ψ are given in the preceding section.

Note that the condition *C* > 0 for the existence of an endemic equilibrium is equivalent to the condition that *R*_0_ > 1. It can be observed that, with values for all other parameters unchanged, *R*_*end*_ increases monotonically as the value for the model parameter *δ* increases. Setting the initial conditions and parameters as in the baseline model, excepting a change in the parameter value of *δ*, simulation results depicting endemic equilibria corresponding to different *R*_*end*_ thresholds are presented in Figures 2, 3, 4.

**Figure 2:**
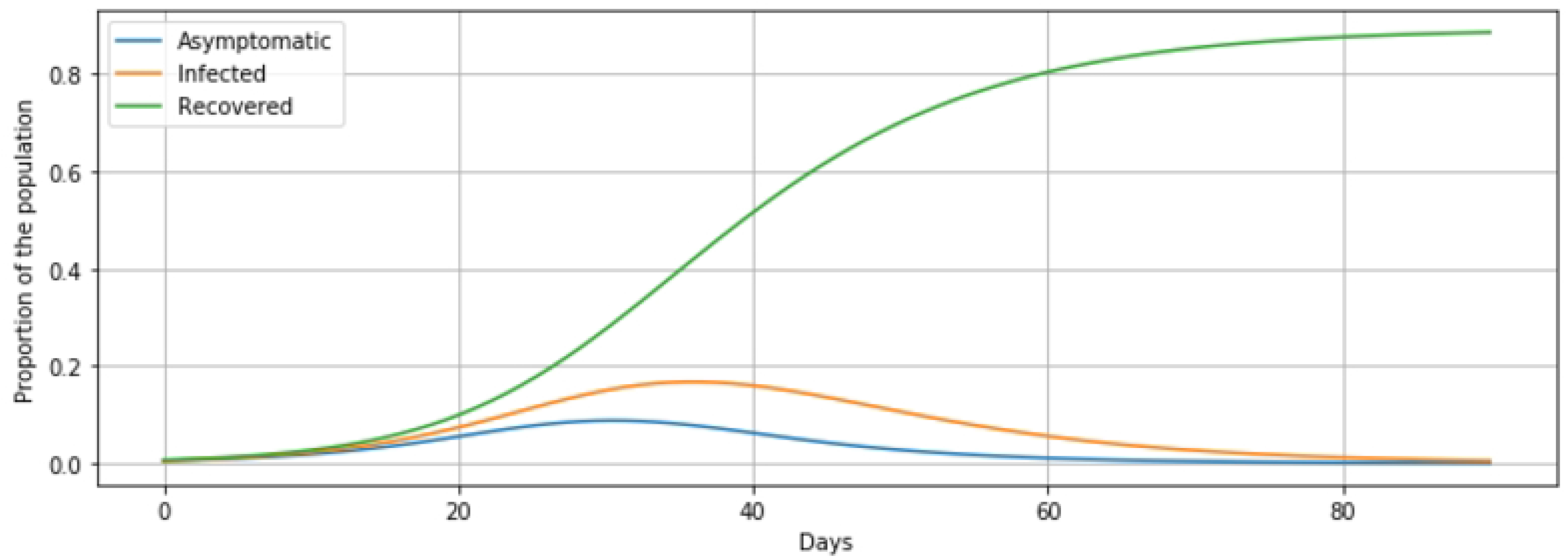
Endemicity with *δ* = 0.001, corresponding to immunity following infection of approximately 2.8 years.

**Figure 3:**
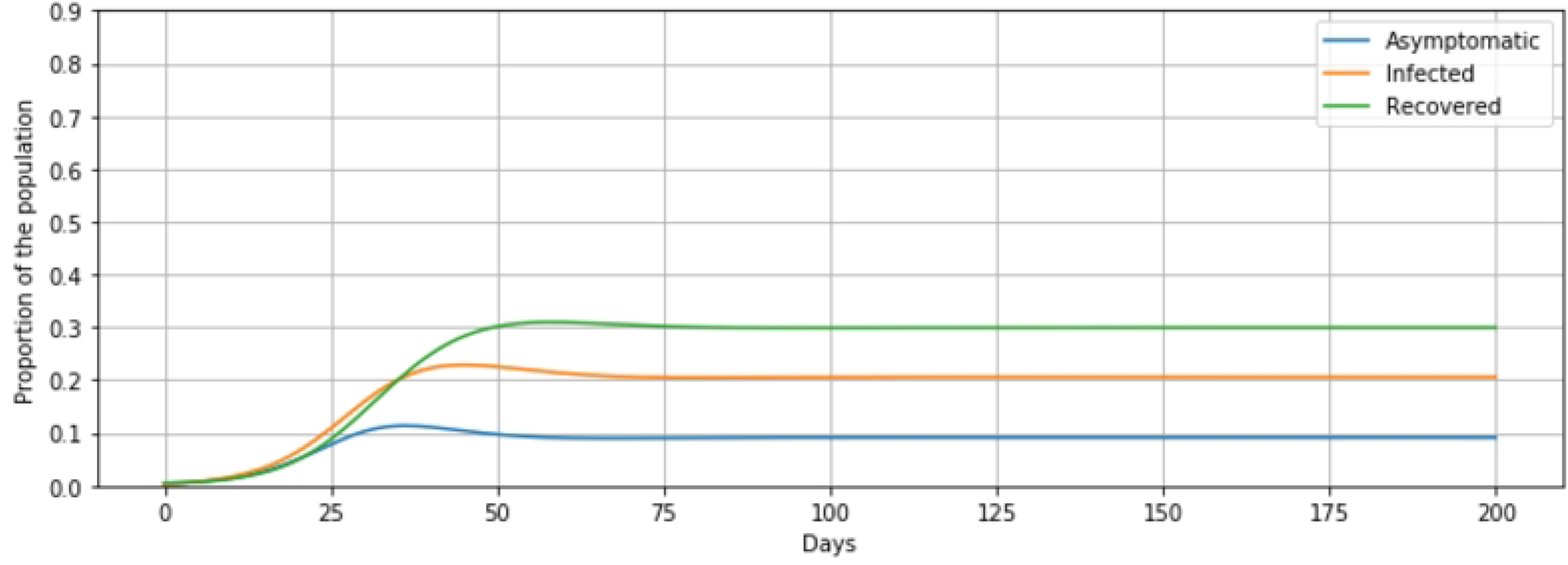
Endemicity with *δ* = 0.01, corresponding to immunity following infection of approximately 3 months.

**Figure 4:**
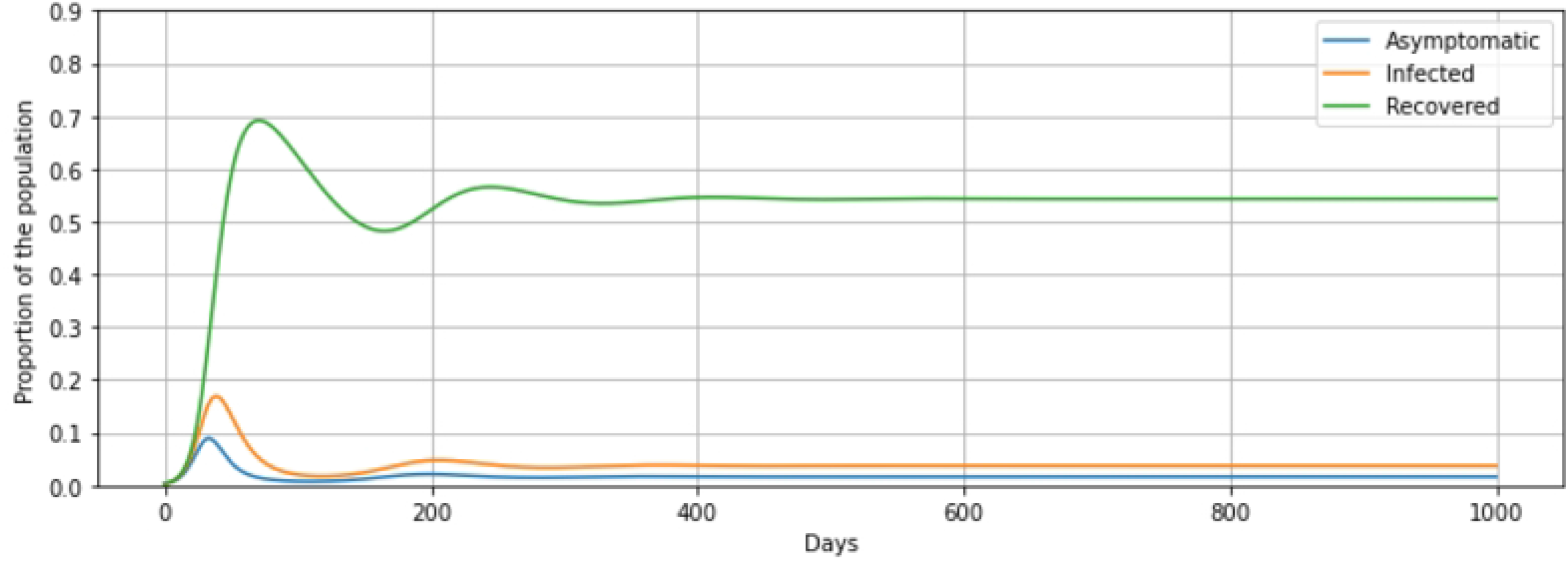
Endemicity with *δ* = 0.1, corresponding to immunity following infection of approximately 10 days.

As the value for *R*_*end*_ increases with increasing *δ* value, the oscillations before reaching the endemic equilibrium have smaller amplitude, although they may have higher frequency. For the models with parameters *δ* = 0.001, *δ* = 0.01, *δ* = 0.1, the first oscillatory dip in the *R* (recovered) subgroup occurs at approximately 850 days, 160 days and 55 days, respectively, and the amplitude differences in the proportions of the recovered subgroups between the first peaks to the following lowest points are approximately 0.45, 0.22, 0.015, respectively. Comparing the population proportions for the endemic equilibria points in these three models, as the value for *δ* increases, the proportion of *R* decreases, whereas proportions for *A* and *I* increase. This observation coincides with the expression of endemic equilibria presented in (10), in Section 2.

### 4.2 Asymptomatic Effects

One major obstacle in the control of COVID-19 has been the challenge of identifying and monitoring individuals in the asymptomatic but infectious subgroup. Herein we explore the impact of the asymptomatic subgroup on the epidemic evolution. We first assume no control actions are imposed on either the asymptomatic or symptomatic infected subgroups, for example, imposing isolation or masking policies. For simplicity, we use the group model (3) with the same parameter values as in our baseline model, which gives a basic reproduction number *R*_0_ ≈ 2.5 from (7).

By setting initial proportions for the *A, I, R* compartments as

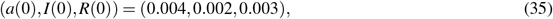

we obtain the 90-day simulation results shown in Fig 5. The population reaches a peak infection level of approximately 25% on day 35. By day 80, approximately 87% of the population has been or is infected.

**Figure 5:**
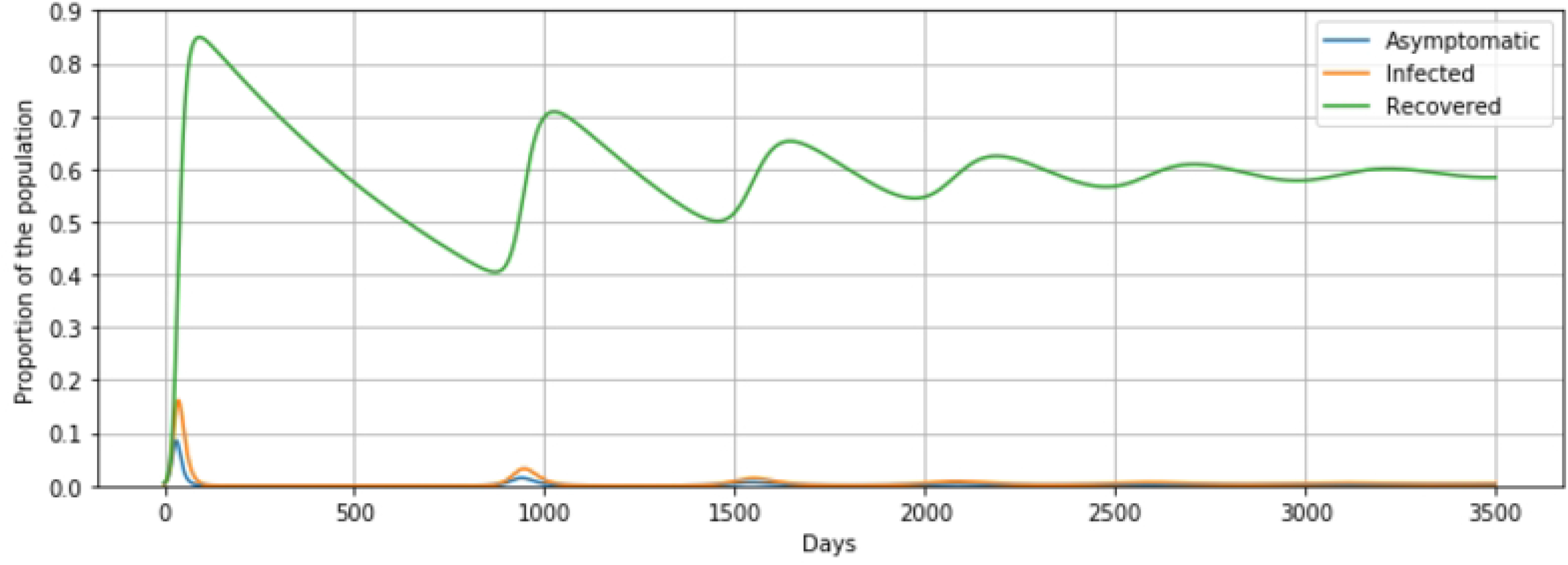
No control policies in effect on either Asymptomatic or Symptomatic Infected subgroups

Next, we implement moderate and stringent isolation policies on only the symptomatic subgroup; this is effected in the simulations by changing the respective infection rate parameters of the subgroups, which we now denote individually by *β*_*A*_ and *β*_*I*_. Imposing isolation policies on a subgroup effectively lowers the corresponding infection rate. The simulations results are shown in Fig 6. We note that with isolation measures on only the symptomatic infected subgroup, the epidemic now progresses more slowly and mildly, as is expected, however there is still substantial infection in the population. The infection peaks at days 60 and 75, respectively, approximately 4 − 6 weeks later than with no control. With moderate isolation policies in effect on the *I* subgroup, the peak infection level is approximately 9% and with strict isolation policies the peak infection level attained is approximately 2.5%. Finally by day 80, the total percentages of the population that have been or are infected is approximately 49% and 17%; with a mortality rate of 2% this corresponds to 98 and 34 deaths, respectively.

**Figure 6:**
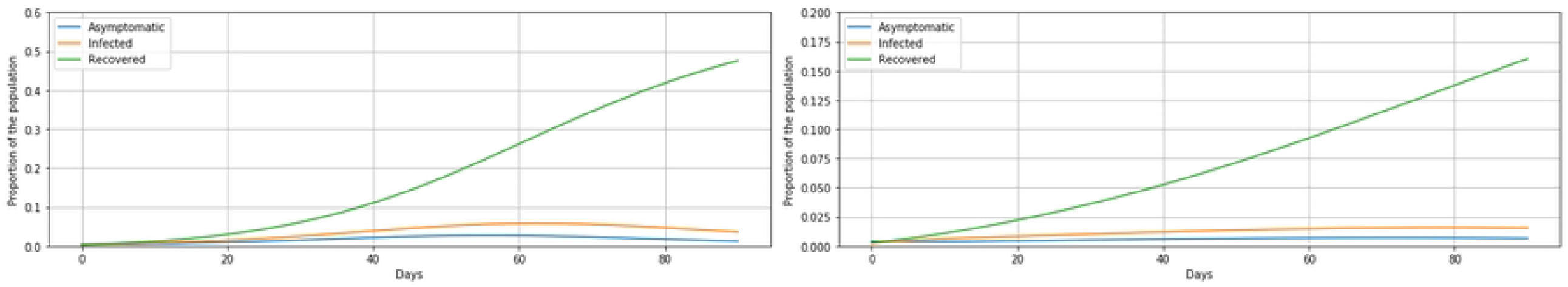
Imposing isolation policies on subgroup *I* (a) Moderate isolation of Symptomatic-Infected subgroup; *β*_*A*_ = 0:25; *β*_*I*_ = 0:11 giving effective *R*_0_ = 1:5 (b) Stringent isolation of Symptomatic-Infected subgroup; *β*_*A*_ = 0:25; *β*_*I*_ = 0:06 giving effective *R*_0_ = 1:2

Alternatively, we consider the situation where Asymptomatic individuals are also identified and isolated, under both moderate and stringent policies, with the results shown in Fig 7.

**Figure 7:**
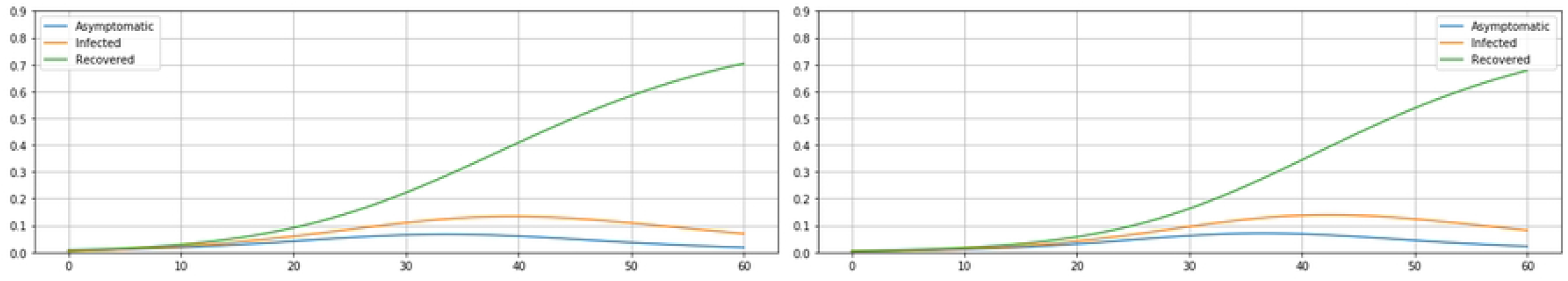
Imposing isolation policies on subgroups *A* and *I* (a) Moderate isolation of both Symptomatic- and Asymptomatic-Infected subgroups; *β*_*A*_ = 0:11; *β*_*I*_ = 0:11 giving effective *R*_0_ = 1:09 (b) Stringent isolation of both Symptomatic- and Asymptomatic-Infected subgroups; *β*_*A*_ = 0:0125; *β*_*I*_ = 0:0125 giving effective *R*_0_ = 0:12

Note that, with only moderate isolation on both Asymptomatic and Symptomatic Infected subgroups (7a), the epidemic is under control within three months. By day 80, approximately 7.7% of the population has been or is infected, corresponding to a total of 770 individuals in a population base of 10, 000; at a 2% mortality rate this corresponds to approximately 15 − 16 deaths as compared to approximately 34 deaths with stringent control imposed on only the Symptomatic Infected subgroup (6b).

An additional perspective to consider is the effective reproduction number under the different isolation policies. Moderate isolation of both Asymptomatic and Symptomatic subgroups (7a) gives an effective *R*_0_ ≈ 1.09, while stringent isolation on just the Symptomatic subgroup (6b) gives an effective *R*_0_ ≈ 1.2.

These simulation results confirm the obvious: identification and isolation of Asymptomatic infected individuals is much more effective in curbing the spread of the epidemic than identification and isolation of only the Symptomatic subgroup. To achieve this goal, either regular extensive mandatory testing policies, or persistent isolation of the whole population, is required.

### 4.3 Network Effects

Here, we evaluate the effect that a more realistic interaction structure has on epidemic spread over a population. We consider the 5-node network introduced earlier, and consider the removal of a small number of edges between nodes, corresponding to there being no interaction between certain subpopulations. We first consider an interconnection network structure with adjacency matrix

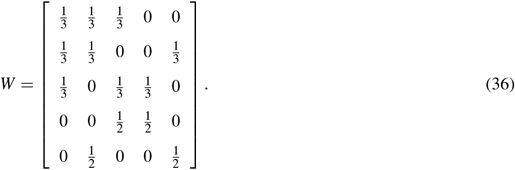

Using the same parameters and initial conditions as in the baseline model, our simulations return results as shown in Fig 8 for subpopulations C and E, for example.

**Figure 8:**
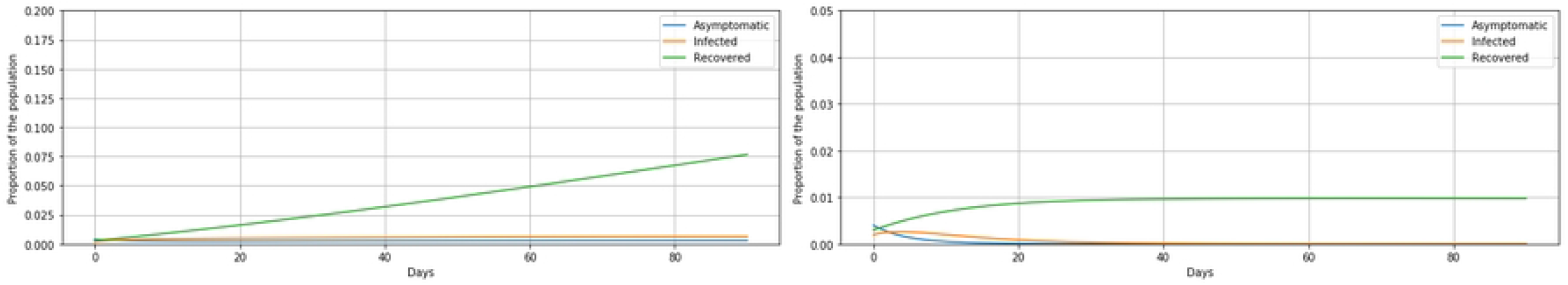
Densely Connected Network Simulation Results (a) Subpopulation C (b) Subpopulation E

With an incomplete network structure, the epidemic spreads more slowly and weakly. Subpopulation C reaches its peak infection level at day 37, and subpopulation E at day 39. By day 60, approximately 83% of area C population and 81% of area E population have been infected. However, in total, approximately 480 fewer individuals over the five areas are infected as compared to the fully connected (i.e., complete) baseline model.

To explore the impact of quarantine and stronger social distancing measures, we further break the full population into 50 smaller subpopulations, and generate a stochastic adjacency matrix with each node only connected to (randomly selected) 20 other nodes out of the total of 50 group nodes. We also generated the initial proportions randomly, i.e., *a*(0), *p*(0), *r*(0), assuming *a*_*i*_(0) ∼ 𝒩 (0.04, 0.005), *p*_*i*_(0) ∼ 𝒩 (0.02, 0.005), *r*_*i*_(0) ∼ 𝒩 (0.03, 0.005), with these values restricted to be non-negative. Randomly selecting 6 of the 50 sub-populations, we present a sample of the simulation results as shown in Fig 9:

**Figure 9:**
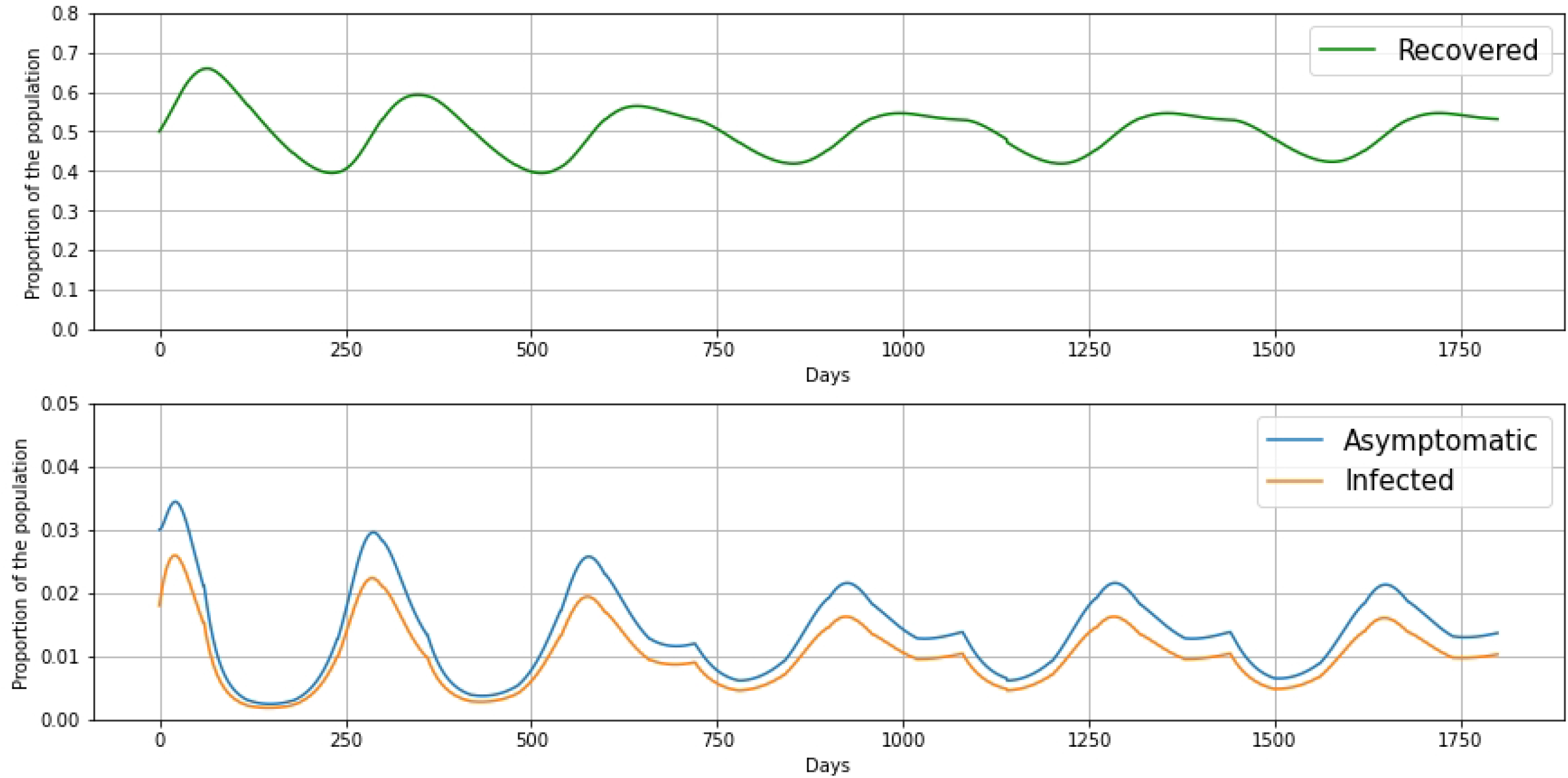
Sparsely Connected Network Simulation Results (a) Subpopulation 4 (b) Subpopulation 22 (c) Subpopulation 28 (d) Subpopulation 32 (e) Subpopulation 34 (f) Subpopulation 48

Note that, with this more extensive isolation structure, the epidemic decays much faster than under the previous densely connected network (Fig 8). Subpopulations 4, 22, 28, 32, 34 and 48, respectively, reach their peak infection levels at days 21, 59, 7, 24, 0 and day 8. Among the six subpopulations in the sample, subpopulation 22 is the most highly infected group. However, overall after 60 days, approximately only 13.6% of the population has been or is infected, which is a reduction of 73.4% of the population compared to the fully connected network (Fig 1), and a reduction of 67.7% compared to the strongly connected network (Fig 8). These simulations confirm that social distancing measures, such as quarantining within each community or family, does serve to slow the spread of the epidemic. From the perspective of the group model, extensive isolation policies do help reduce the group transmission rate for person-to-person contact, although these do not completely halt the disease spread.

We also investigate the impacts of different underlying network structures on the stability of the DFE and endemic equilibria. As shown in (23) in Section 2, the bound provided on the eigenvalues of the *q*-scaled adjacency matrix *W* by the eigenvalues of a matrix generated by diagonal block entries of ratios of recovery and transition rates (symptoms onsetting) to infection rates gives a sufficient condition for the stability of the DFE. We consider the equilibria for the full population under different interconnection network structures. Evaluating endemic equilibria, for all *i* ∈ [*n*], we set the homogeneous spread parameter values to

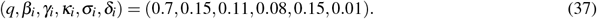

The convergence results of the networked models (4) with a 5-node densely connected network (36), a 50-node fully connected network and a 50-node sparsely connected network are illustrated in Fig 10, yielding the simulation results shown over 700 days.

**Figure 10:**
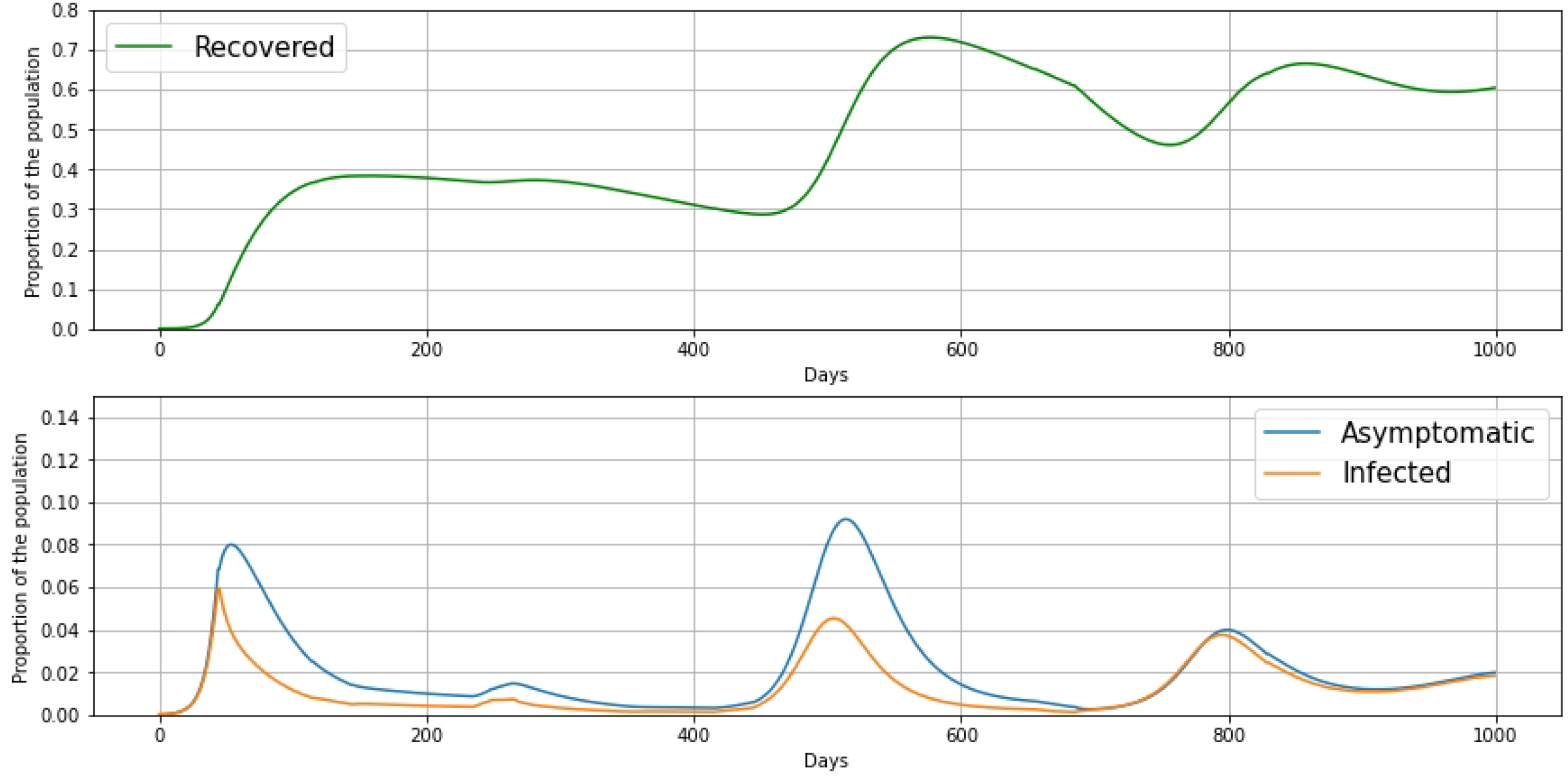
DFE convergence for N-SAIRS models (a) Fully connected network (b) Densely connected network (c) Sparsely connected network

On one hand, the maximum eigenvalue of the fully connected graph (10a) and densely connected graph (10b) are both approximately 1.04. This value is larger than the minimum eigenvalue of the parameters matrix, which is approximately 0.49. In both network structures, the systems converge to the endemic equilibrium. On the other hand, the maximum eigenvalue of the sparsely connected graph (10c), which is approximately 0.25, is smaller than the minimum eigenvalue of the parameters matrix. With this network structure, the system converges to the DFE. These simulation results serve as examples of the condition derived in (23). Note that (23) is not a necessary condition for convergence to the DFE. That is, violation of this condition does not guarantee that the system will converge to an endemic equilibrium.

### 4.4 A Multi-stage Simulation

In addition to non-pharmaceutical measures, such as mask policies and social distancing, pharmaceutical measures such as vaccinations and treatment of symptoms clearly also play important roles in the control of epidemic spread processes. With the increasingly availability of vaccines for COVID-19 since March 2021, the number of daily new cases of COVID-19 has dropped, even with less strict social distancing amongst populations [54, 55]. However, the overall effect of these vaccinations may be compromised by virus mutations [56, 57]. In this section, taking both vaccines and virus mutations into consideration, we present a multi-stage group SAIRS simulation representing the evolution of COVID-19 in Illinois in Fig 11; the relevant timeline is also given.

**Figure 11:**
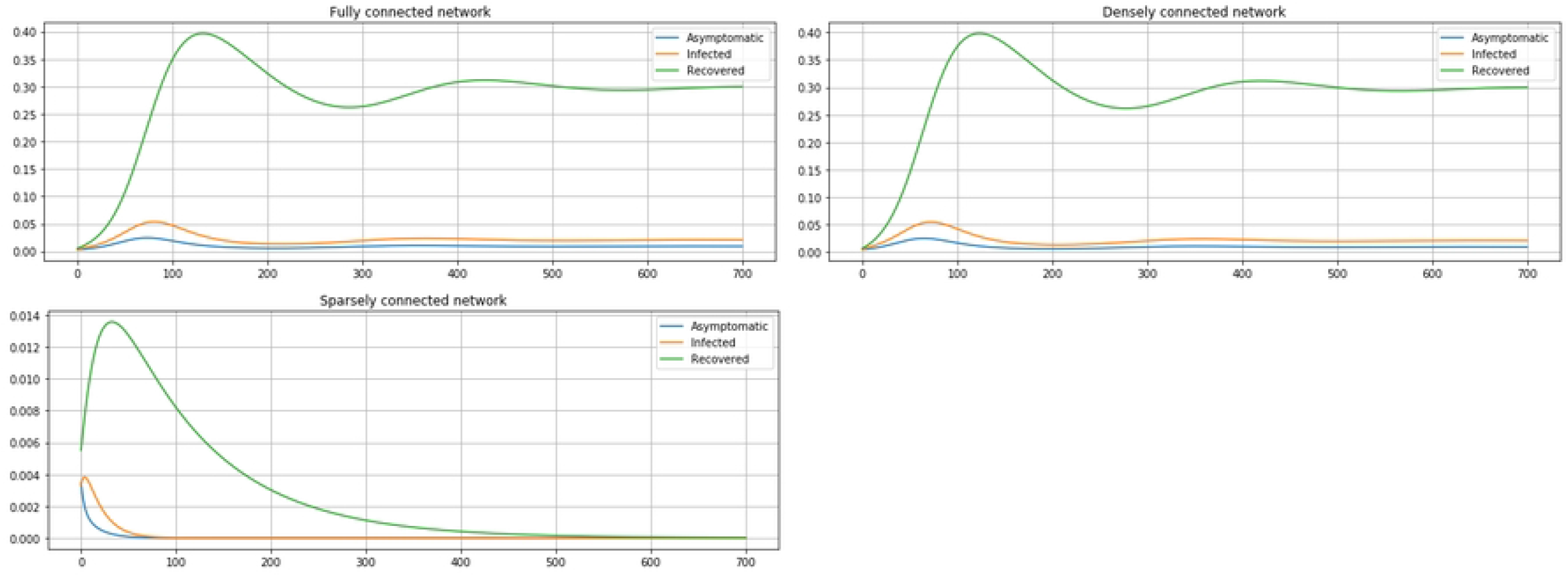
Multi-stage (3-year) SAIRS process with Vaccinations and Virus Mutations

*Stage* 1 : Rapid Spread (45 *days* : *Day* 1 − *Day* 45)

*Stage* 2 : Flattening: *Lockdown* (70 *days* : *Day* 46 − *Day* 115)

*Stage* 3 : Restoration: *Social distancing, masking, reduced indoor capacities* (135 *days* : *Day* 116 − *Day* 250)

*Stage* 4 : Re-enacted Restrictions: 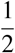 *o f all regions* (15 *days* : *Day* 251 *Day* 265)

*Stage* 5 : Re-enacted Restrictions: *all regions* (90 *days* : *Day* 266 − *Day* 355)

*Stage* 6 : Vaccination rollout: (90 *days* : *Day* 356 − *Day* 445)

*Stage* 7 : Variant 1 Emergence: (210 *days* : *Day* 446 − *Day* 655)

*Stage* 8 : Booster rollout: (30 *days* : *Day* 656 − *Day* 685)

*Stage* 9 : Variant 2 Emergence: (145 *days* : *Day* 686 − *Day* 830)

*Stage* 10 : Social Distancing Lifted, Waning Vaccine Immunity: (170 *days* : *Day* 831 − *Day* 1000)

We have again considered the baseline group model

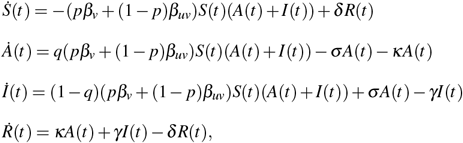

*p* is the vaccination level, and *β*_*v*_, *β*_*uv*_, respectively, are the transmission rates between infected and non-infected individuals who have or have not been vaccinated, respectively. Assuming that the immunity individuals gain after infection or vaccination lasts one year, we set the baseline model parameter values based on COVID-19 literature for the United States up to June 2021 as

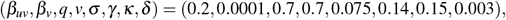

Different *β*_*uv*_ values are implemented throughout the simulation for different levels of social distancing. In Stage 6, the vaccination level *p* is set to increase at the beginning of each sub-stage. Specifically, *p* = 0.2 for day 1-day 30; *p* = 0.4 for day 31-day 60, *p* = 0.5 for day 61-day 90, in reference to the first day of vaccination rollout. In Stage 7, both *β*_*uv*_ and *β*_*v*_ are set to be higher than the original baseline transmission rates due to the highly contagious nature of the new variant, and the receding immunity provided by the vaccines as time passes.

As shown in Figure 11, the lockdown in Stage 2 effectively mitigates the spread of the epidemic, whereas the relaxation of social distancing policies results in a slight surge in infection rates starting from approximately day 240. This coincides with the network results previously presented in Section 4.3. Vaccination in Stage 6 successfully mitigates the spread of the epidemic, however, a new surge arises with the emergence of new variants starting from approximately day 450, which attains a peak infection level of approximately 13.5% on day 510. To reinforce the effect of the first round of vaccines, in Stage 8 booster shots are provided, which facilitate the mitigation of the epidemic. The effectiveness of these booster shots are again undermined as a more contagious variant of the virus emerges on day 685. Despite being highly contagious, this new variant appears to result in less severe symptoms among vaccinated individuals, which results in faster recoveries compared to the previous virus strains. As the immunity gained from vaccines recedes over time and social distancing policies are lifted, individuals become more exposed to new variants and therefore have higher chances of being infected or reinfected, resulting in another surge in the infection level during stage 10.

Further, we simulated a five-year glance into the future where the epidemic of COVID-19 potentially becomes endemic, and updated booster vaccines are provided annually, similar to current influenza practices; this is shown in Fig 12.

**Figure 12:**
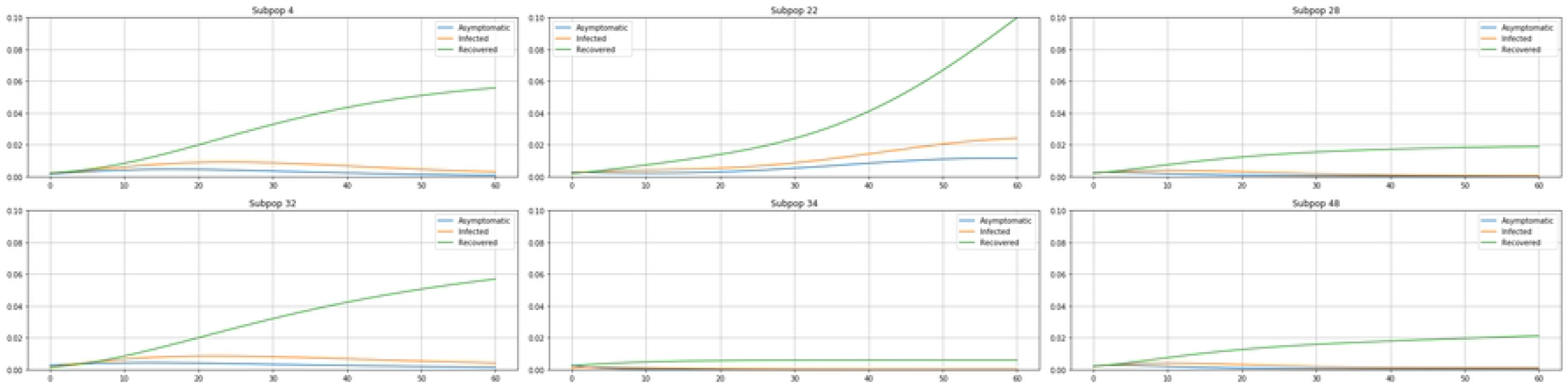
Endemic SAIRS with Annual Vaccinations

We have again updated model parameter values as

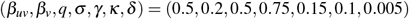

based on the COVID-19 literature up to September 2022 (e.g. [58–61]), where Omicron (B.1.1.529) is the main variant of the endemic epidemic. We also assume the effectiveness of the annual vaccines decreases by 10% every two months based on [62]. This corresponds to 100% effectiveness during the first two months following vaccination, and 50% effectiveness over the last two months until getting the next annual vaccination. We set the initial proportions for the *A, I, R* compartments as

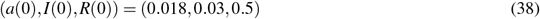

based on the proportions at the end of Multi-stage simulation (Fig 11) and [63]. Fig 12 shows periodic oscillations in both the recovered population (*R*: attaining local peaks at day 64, day 347, day 642, day 996, day 1356 and day 1721) and the infected populations (*A* + *I*: attaining local peaks at day 21, day 287, day 576, day 924, day 1284 and day 1648), for which the frequency and amplitude decreases over time until eventually reaching the endemic equilibria. This observed epidemic behavior coincides with the behavior of the endemic evolution as simulated in Figures 2,3,4.

## 5 Conclusions and Future Work

In this paper, we have briefly reviewed classical epidemiological compartment models, with a focus on a new SAIR(S) model that emphasizes the role played by the Asymptomatic-infected subpopulation. We presented continuous-time, discrete-time, and networked versions of the SAIR(S) model, and discuss their corresponding equilibria and stability properties. We have noted the use of Nesterov’s Next-Day Law and a basic least-squares approach for model parameter estimation, and conducted initial parameter estimation for COVID-19 using publicly available data from Champaign County, Illinois. Furthermore, we completed simulations of both group and networked models investigating the impact of isolating subpopulations, highlighting the crucial role of the Asymptomatic subgroup in the control of epidemic evolution, and exploring long-term endemicity conditions.

In the estimation process, we have encountered many challenges, most significantly biased testing data and the lack of explicit information on the asymptomatic infected population. Our ongoing efforts include pursuing more complete endemic equilibria analyses for the N-SAIR(S) model and investigating approaches for model estimation under non-random and missing sample data sets, for example as described in [64]. We are further investigating Bayesian statistical methods for estimating true prevalence of epidemics given biased information for apparent prevalence [65].

## Data Availability

All datasets are available from the figshare database DOI: 10.6084/m9.figshare.21902580 10.6084/m9.figshare.21903072

https://doi.org/10.6084/m9.figshare.21902580

https://doi.org/10.6084/m9.figshare.21903072

## Acknowledgments

The authors would like to thank Dr. Joseph Kim, M.D., Ph.D., for many useful and interesting discussions, and for providing informative references.

## References

[1] Yin Q, Shi T, Dong C, Yan Z. The impact of contact patterns on epidemic dynamics. PLOS ONE. 2017.

[2] Oliva G, Schlueter M, Munetomo M, Scala A. Dynamical intervention planning against COVID-19-like epidemics. PLOS ONE. 2022.

[3] Kermack WO, McKendrick AG. Contributions to the mathematical theory of epidemics. II. The problem of endemicity. Proceedings of the Royal Society A. 1932;138(834):55–83.

[4] Bernoulli D. Essai d’une nouvelle analyse de la mortalité causée par la petite vérole et des avantages de l’inoculation pour la prévenir. Histoire de l’Acad Roy Sci avec Mém des Math et Phys and Mém. 1760:1–45.

[5] Scarpino SV, Petri G. On the predictability of infectious disease outbreaks. Nature communications. 2019;10.

[6] Funk S, Gilad E, Watkins C, Jansen VAA. The spread of awareness and its impact on epidemic outbreaks. Proceedings of The National Academy of Sciences. 2009;2009:6872–6877.

[7] Granell C, Gómez S, Arenas A. Dynamical Interplay between Awareness and Epidemic Spreading in Multiplex Networks. Phys Rev Lett. 2013 Sep;111:128701. Available from: http://link.aps.org/doi/10.1103/PhysRevLett.111.128701.

[8] Paarporn K, Eksin C, Weitz JS, Shamma JS. Networked SIS Epidemics with Awareness. IEEE Trans Computational Social Systems. 2017;4(3):93–103.

[9] Liu J, Paré PE. Nedić A, Beck CL, Başar T. On a continuous-time multi-group bi-virus model with human awareness. In: Proceedings of The IEEE Conference on Decision and Control; 2017. p. 4124–4129.

[10] Xu S, Lu W, Zhan Z. A Stochastic Model of Multivirus Dynamics. IEEE Transactions on Dependable and Secure Computing. 2012;9(1):30–45.

[11] Paré PE, Liu J, Beck CL, Nedić A, Başar T. Multi-Competitive Viruses over Static and Time–Varying Networks. In: Proceedings of American Control Conference; 2017. p. 1685–1690.

[12] Paré PE, Liu J, Beck CL, Nedić A, Basar T. Multi-Competitive Viruses over Time-Varying Networks with Mutations and human Awareness. Automatica. 2021;123.

[13] Li G, Zhang Y. Dynamic behaviors of a modified SIR model in epidemic diseases using nonlinear incidence and recovery rates. PLOS ONE. 2017.

[14] Kephart JO, White SR. Directed-graph epidemiological models of computer viruses. In: IEEE Symposium on Security and Privacy; 1991. p. 343–361.

[15] Newman ME. Spread of epidemic disease on networks. Phys Rev E Stat Nonlin Soft Matter Phys. 2002 06;66.

[16] Ganesh A, Massouli L, Towsley D. The effect of network topology on the spread of epidemics. In: 24th Annual Joint Conference of the IEEE Computer and Communications Societies. vol. 2; 2005. p. 1455–1466.

[17] Draief M, Massoulie’ L. Epidemics and rumours in complex networks. Cambridge University Press; 2010.

[18] Pastor-Satorras R, Castellano C, Van Mieghem P, Vespignani A. Epidemic processes in complex networks. Reviews of Modern Physics. 2015;87(3):925.

[19] Nowzari C, Preciado VM, Pappas GJ. Analysis and control of epidemics: A survey of spreading processes on complex networks. IEEE Control Systems Magazine. 2016;(1):26–46.

[20] Pare P, Beck CL, Başar T. Modeling, Estimation, and Analysis of Epidemics over Networks: An Overview. Annual Reviews in Control. 2020;2020:345–360.

[21] Zhao D, Wang L, Li S, Wang Z, Wang L, Gao B. Immunization of Epidemics in Multiplex Networks. PLOS ONE. 2014.

[22] Feller W. On the Integro-Differential Equations of Purely Discontinuous Markoff Processes. Transactions of the American Mathematical Society. 1940;48(3):488–515.

[23] Kurtz TG. The Central Limit Theorem for Markov Chains. The Annals of Probability. 1981;9(4):557–560.

[24] Mieghem PV, Omic J, Kooij R. Virus spread in networks. IEEE/ACM Transactions on Networking. 2009;(1):62– 68.

[25] Chatterjee S, Durrett R. Contact processes on random graphs with power law degree distributions have critical value 0. The Annals of Probability. 2009;(6):2332–2356.

[26] Fall A, Iggidr A, Sallet G, Tewa JJ. Epidemiological models and Lyapunov functions. Mathematical Modelling of Natural Phenomena. 2007;2(1):62–83.

[27] Paré PE, Beck CL, Nedić A. Epidemic Processes over Time-Varying Networks. IEEE Transactions on Control over Network Systems. 2018;(3):1322–1334.

[28] Ahn HJ, Hassibi B. Global dynamics of epidemic spread over complex networks. In: Proceedings of the IEEE Conference on Decision and Control; 2013. p. 4579–4585.

[29] Wang Y, Chakrabarti D, Wang C, Faloutsos C. Epidemic spreading in real networks: an eigenvalue viewpoint. In: Proceedings of the 22nd International Symposium on Reliable Distributed Systems; 2003. p. 25–34.

[30] Khanafer A, Başar T, Gharesifard B. Stability properties of infected networks with low curing rates. In: Proceedings of the American Control Conference; 2014. p. 3579–3584.

[31] Khanafer A, Başar T, Gharesifard B. Stability properties of infection diffusion dynamics over directed networks. In: Proceedings of the IEEE Conference on Decision and Control; 2014. p. 6215–6220.

[32] Nowzari C, Preciado VM, Pappas GJ. Stability analysis of generalized epidemic models over directed networks. In: Proceedings of the IEEE Conference on Decision and Control; 2014. p. 6197–6202.

[33] Health Care Engineering Systems Center U. COVID-19 Virtual Summit; 2020. April 6.

[34] NeTs Community N, the Ohio State University. First Call to Arms Workshop; 2020. April 13.

[35] Bi X, Beck CL. On the Role of Asymptomatic Carriers in Epidemic Spread Processess. arXiv. 2021 March. ArXiv:2103.11411.

[36] Grunnill M. An exploration of the role of asymptomatic infections in the epidemiology of dengue viruses through susceptible, asymptomatic, infected and recovered (SAIR) models. Journal of Theoretical Biology. 2018;2018:195–204.

[37] Zhu L, Wang B. Stability analysis of a SAIR rumor spreading model with control strategies in online social networks. Information Science. 2020;526.

[38] Hota A, Sneh T, Gupta K. Impacts of Game-Theoretic Activation on Epidemic Spread over Dynamical Networks. arXiv. 2020 Nov. ArXiv2011.00445v1 [physics.soc-ph].

[39] Dobrovolny HM. Modeling the role of asymptomatics in infection spread with application to SARS-CoV-2. PLOS ONE. 2020.

[40] Paiva HM, Afonso RJM, Oliveira IL, Garcia GF. A data-driven model to describe and forecast the dynamics of COVID-19 transmission. PLOS ONE. 2020.

[41] Stella L, Martinez AP, Bauso D, Colanari P. The Role of Asymptomatic Individuals in the COVID-19 Pandemic via Complex Networks. arXiv. 2020 Sept. ArXiv:2009.03649v1 [physics.soc-ph].

[42] Khalil HK. Nonlinear Systems. Prentice Hall; 2002.

[43] Paré PE, Liu J, Beck CL, Kirwan BE, Başar T. Analysis, Identification, and Validation of Discrete-Time Epidemic Processes. IEEE Transactions on Control Systems Technology. 2019;28(1):79–93.

[44] Nesterov Y. Online prediction of COVID19 dynamics.Belgian case study. LIDAM Discussion Papers CORE. 2020.

[45] Bi X, Dekhterman S, Beck CL. Champaign County SARS-CoV-2 Test Site Data. figshare; 2023. http:///doi.org/10.6084/m9.figshare.21902580.

[46] Peoria SARS-CoV-2 clinic data. figshare; 2023. https://doi.org/10.6084/m9.figshare.21903072.

[47] Hoff V. Estimation of Hidden Carriers of Infectious Diseases [Master’s thesis]. University of Iliinois at Urbana-Champaign; 2022. Available from: http://www.ideals.illinois.edu/items/124464.

[48] Lauer SA, Grantz KH, Bi Q, Jones FK, Zheng Q, Meredith HR, et al. The Incubation Period of Coronavirus Disease 2019 (COVID-19) From Publicly Reported Confirmed Cases: Estimation and Application. Annals of Internal Medicine. 2020.

[49] Oran DP, E J Topol M. Prevalence of Asymptomatic SARS-CoV-2 Infection; A Narrative Review. Annals of Internal Medicine. 2020.

[50] Ling Y, Xu S, Lin Y, Tian D, Zhu Z, Dai F, et al. Persistence and clearance of viral RNA in 2019 novel coronavirus disease rehabilitation patients. Chinese Medical Journal. 2020;133.

[51] Li Q, Guan X, Wu P, Wang X, Zhou L, Tong Y, et al. Early Transmission Dynamics in Wuhan, China, of Novel Coronavirus–Infected Pneumonia. N Engl J Med. 2020;382.

[52] Arons MM, Hatfield KM, Reddy SC, Kimball A, James A, Jacobs JR, et al. Presymptomatic SARS-CoV-2 Infections and Transmission in a Skilled Nursing Facility. N Engl J Med. 2020;382.

[53] Paré PE, Liu J, Beck CL, Nedić A, Başar T. Multi-Competitive Viruses over Time–Varying Networks with Mutations and Human Awareness. 2020.

[54] Moghadas SM, Vilches TN, Zhang K, et al. The impact of vaccination on COVID-19 outbreaks in the United States. Clinical infectious diseases : an official publication of the Infectious Diseases Society of America. 2021 Jan.

[55] Evine-Tiefenbrun M, Yelin I, Katz R, et al. Initial report of decreased SARS-CoV-2 viral load after inoculation with the BNT162b2 vaccine. Nature Medicine. 2021 March.

[56] Bernal JL, et al. Effectiveness of Covid-19 Vaccines against the B.1.617.2 (Delta) Variant. The New England Journal of Medicine. 2021 July.

[57] Bian L, Gao F, Zhang J, et al. Effects of SARS-CoV-2 variants on vaccine efficacy and response strategies. Expert review of vaccines. 2021 April.

[58] Centers for Disease Control and Prevention. SARS-CoV-2 B.1.1.529 (Omicron) Variant Transmission Within Households — Four U.S. Jurisdictions, November 2021–February 2022. Morbidity and Mortality Weekly Report. 2022 03.

[59] Liu Y, Rocklöv J. The effective reproductive number of the Omicron variant of SARS-CoV-2 is several times relative to Delta. Journal of Travel Medicine. 2022 05.

[60] Pulliam JRC, Schalkwyk CV, Govender N, Gottberg AV, Cohen C, Groome MJ, et al. Increased risk of SARS-CoV-2 reinfection associated with emergence of Omicron in South Africa. Science. 2022 05.

[61] Moghadas SM, Vilches TN, Zhang K, Wells CR, Shoukat A, Singer BH, et al. The Impact of Vaccination on Coronavirus Disease 2019 (COVID-19) Outbreaks in the United States. Clinical Infectious Diseases. 2021 01;73(12):2257–2264.

[62] Feikin DR, Higdon MM, Abu-Raddad LJ, Andrews N, Araos R, Goldberg Y, et al. Duration of effectiveness of vaccines against SARS-CoV-2 infection and COVID-19 disease: results of a systematic review and metaregression. The lancet. 2022 05.

[63] Centers for Disease Control and Prevention. Nationwide COVID-19 Infection-Induced Antibody Seroprevalence (Commercial laboratories); 2022. Data retrieved from COVID Data Tracker.

[64] Copas JB, Li HG. Inference for non-random samples. Journal of the Royal Statistical Society. 1997;59(1):55–95.

[65] Bi X, Miehling E, Beck CL, Başar T. Approximate Testing in Uncertain Epidemic Processes. In: Proceedings of the IEEE Conference on Decision and Control; 2022..

